# Assessing the Diagnostic Accuracy of Brain MR Spectroscopy for Various Neurologic Pathologies: A Comparative Study with Final Clinical Diagnoses in Patients Referred to Shiraz University Hospitals (2021–2024)

**DOI:** 10.1101/2025.07.06.25330968

**Authors:** Seyedeh Zahra Mousavi, Fatemeh Yarmahmoodi

**Affiliations:** Medical Imaging Research center, Department of Radiology, Shiraz University of Medical Sciences, Shiraz, Iran

**Author notes:** Corresponding author Fatemeh Yarmahmoodi, Medical Imaging Research center, Department of Radiology, Shiraz University of Medical Sciences, Shiraz, Iran.

**Keywords:** magnetic resonance spectroscopy, magnetic resonance imaging, neurologic disorders, brain tumor, demyelinating disease

## Abstract

**Introduction:** Magnetic resonance spectroscopy (MRS) of the brain offers significant potential for monitoring metabolic alterations associated with neurological pathologies. However, to date, its application has predominantly been limited to research settings and a select number of clinical centers. This study aimed to assess the diagnostic accuracy of brain MRS in detecting various neurological diseases.

**Methods:** Brain MRS examinations conducted at public hospitals affiliated with Shiraz University, Iran, from 2021 to 2024 were retrospectively collected. The findings, as reported by attending radiologists, were compared with the final clinical or histopathological diagnoses of the patients. Diagnostic performance metrics, including sensitivity, specificity, positive predictive value, negative predictive value, and overall accuracy, were calculated.

**Results:** Brain MRS correctly identified the pathology in 61% of cases. The technique demonstrated high specificity, positive predictive value, negative predictive value, and overall accuracy—each exceeding 90%—across a range of brain pathologies. These included both primary and secondary tumoral processes, as well as non-tumoral conditions such as pyogenic brain abscess, toxoplasmosis, demyelinating diseases, and encephalitis. However, sensitivity varied considerably, ranging from 33% to 100%, with the lowest sensitivity observed in cases of pyogenic brain abscesses, brain hemorrhages, and central nervous system (CNS) vasculitis.

**Conclusion:** This study indicates that certain neurological pathologies may exhibit atypical spectroscopic features that are not yet fully characterized. The findings underscore the necessity for further research involving larger patient populations to better delineate the spectroscopic patterns associated with these conditions.

## INTRODUCTION

Magnetic Resonance Spectroscopy (MRS) is a sophisticated imaging modality that integrates the molecular differentiation capabilities of nuclear magnetic resonance with the spatial and anatomical resolution of Magnetic Resonance Imaging (MRI). This technique enables the noninvasive assessment of tissue biochemical composition, facilitating the identification of various physiological and pathological processes. Notably, MRS does not require the use of contrast agents and does not expose patients to ionizing radiation, representing a significant clinical advantage (1–3).

MRS can be acquired in either single-voxel or multi-voxel modes. Single-voxel MRS involves the acquisition of spectral data from a single, localized volume. This approach offers shorter acquisition time and is suitable for rapid evaluation of most brain regions. In contrast, multi-voxel MRS allows simultaneous collection of spectra from multiple voxels, enabling the mapping of regional metabolite distributions and the generation of metabolic images (4). While single-voxel MRS is relatively quick and straightforward to perform, its limitation lies in the inability to assess spatial heterogeneity of metabolites across different brain regions. Consequently, most clinical studies using this method focus on one or two regions. Multi-voxel MRS, although more time-consuming, provides comprehensive spatial information and can evaluate multiple regions simultaneously.

As mentioned, MRS offers a noninvasive, quantitative profile of tissue metabolites, which has proven valuable in the clinical management of various brain diseases. A substantial body of research has explored its applications in neuro-oncology, particularly in the diagnosis and characterization of brain tumors. While some MRS-based protocols have been integrated into routine clinical practice, others remain investigational. Clinically, MRS is instrumental in differentiating primary brain tumors from mimicking conditions such as demyelinating diseases, lymphoma, or infections. Additionally, the molecular profiles obtained via MRS can distinguish between high-grade and low-grade tumors, aiding in prognostication and treatment planning. The technique is also critical for monitoring therapeutic response and detecting tumor recurrence post-treatment, including differentiating radiation-induced necrosis from tumor progression (5, 6).

Despite these advantages, several limitations hinder the widespread clinical adoption of MRS. The overlapping spectroscopic features among different pathologies pose a significant challenge; although each condition exhibits characteristic metabolic patterns, these are often only present in 60–80% of cases. The absence of definitive imaging biomarkers reduces the confidence in MRS findings, limiting its routine diagnostic utility (6).

In particular, differentiating brain metastases, especially solitary lesions, from primary brain tumors remains challenging. Both can present as enhancing masses with surrounding edema on T2/FLAIR images. MRS typically shows increased choline (Cho) and decreased N-acetylaspartate (NAA) levels in both metastases and gliomas, complicating differential diagnosis (7-9). Demyelinating lesions may mimic primary tumors on conventional imaging, and studies have shown variable results regarding MRS’s discriminative capacity; for example, a Cho/NAA ratio exceeding 1.72 has been suggested to favor high-grade gliomas over demyelinating diseases (10), though other research reports no significant differences in ratios such as Cho/Cr between these entities (11). Pyogenic brain abscesses can also be mistaken for neoplasms; classic MRS findings include peaks of amino acids such as valine, alanine, leucine, acetate, and succinate. However, sensitivity and specificity remain limited, with amino acids detected in approximately 80% of abscesses but only moderate diagnostic accuracy (12,13).

The overlapping spectroscopic signatures among astrocytoma, lymphoma, demyelinating diseases, and abscesses underscore the difficulty of relying solely on MRS for definitive diagnosis. Accurate differentiation often requires integrating MRS findings with MRI features and clinical context. Further research is necessary to evaluate the diagnostic performance of MRS in these complex cases and to establish reliable biomarkers that can enhance its clinical utility.

Also, the adoption of MRS into routine clinical workflows has been impeded by technical challenges, including lengthy acquisition times, limited spatial resolution, variable data quality, and the paucity of advanced analysis software. Additionally, there remains a limited awareness within the clinical community regarding the potential impact of MRS on patient management (14, 15).

Building on this understanding, the present study aims to analyze brain MRS findings in a diverse patient cohort, comparing spectroscopic results and differential diagnoses with final clinical diagnoses. The goal is to assess the diagnostic accuracy of MRS across various neurological conditions and to identify areas where technological and methodological advancements are needed to facilitate the integration of advanced MRS techniques into clinical practice. This study is among the first to encompass a broad spectrum of brain pathologies, including less-investigated disorders, providing a comprehensive evaluation of MRS’s diagnostic performance.

## METHODOLOGY

This study will employ a cross-sectional observational design to evaluate the diagnostic accuracy of MRS in differentiating brain lesions and pathologies. It will be conducted at Shiraz University of Medical Sciences, adhering to all ethical guidelines and obtaining necessary institutional approval prior to data recruitment.

The study will include all patients who have undergone brain MRS at public hospitals affiliated with Shiraz University of Medical Sciences from January 2021 to December 2024. Inclusion criteria stipulate that all MRS studies must be of sufficient quality without significant artifacts to allow for interpretation, accompanied by a documented report. Studies conducted solely for research purposes will be excluded. Participants must possess a definitive clinical diagnosis established by the treating medical team.

Results of the MRS have been reported by experienced radiologists. The findings, interpretations, and proposed differential diagnoses will be systematically documented using a structured format devised by the authors. Clinical records for all patients included in the study will be retrieved from the hospital’s archives within the Hospital Information System (HIS), encompassing pathology reports, surgical histories, clinical courses, progress notes, and follow-up documentation. The final clinical diagnoses determined by histopathological examination, clinical follow-up, or imaging follow-up will be compared with the findings and differentials obtained from MRS.

To assess the diagnostic performance of MRS for various neurological pathologies, the following metrics will be calculated: sensitivity, specificity, positive predictive value (PPV), negative predictive value (NPV), and diagnostic accuracy, providing valuable insights into the diagnostic efficacy of MRS for different neurologic disorders.

## RESULTS

A total of 182 MRS studies were conducted from January 2021 to December 2024 at public hospitals affiliated with Shiraz University of Medical Sciences. Images acquired for research purposes, along with those of poor quality or containing significant artifacts, were excluded from the analysis. Clinical diagnoses for 146 cases were obtained from hospital admission files or pathology reports through the HIS, while documentation for 36 cases was unavailable. These patients either were not admitted to the hospital, resulting in missing files, or had passed away without providing contact information. Consequently, 146 MRS studies were included in the final analysis, with findings compared against the patients’ definitive clinical diagnoses.

Among the 146 MRS images, 33 studies provided multiple differential diagnoses without identifying the most probable entity, or the MRS patterns were non-specific, limiting their utility for referring physicians. In the remaining 113 cases, MRS diagnoses aligned with the clinical diagnoses in 89 instances, while discrepancies occurred in 24 cases. Thus, 61% of the 146 MRS studies successfully identified the correct final diagnosis. The data were further stratified based on the final clinical diagnoses to evaluate the performance of MRS for each neurological pathology. Sensitivity, specificity, PPV, NPV, and accuracy of MRS for each pathological entity are summarized in Table 1.

**Table 1.** Sensitivity, specificity, PPV, NPV, and accuracy of MRS for different neurologic pathologies.

|  | sensitivity | specificity | PPV | NPV | accuracy |
| --- | --- | --- | --- | --- | --- |
| Primary brain tumor | 74% | 97% | 92% | 88% | 89% |
| Brain metastasis | 50% | 99% | 67% | 99% | 98% |
| Brain lymphoma | 67% | 98% | 57% | 99% | 97% |
| Recurrence of previously treated tumor | 92% | 100% | 100% | 99% | 99% |
| Demyelinating disease | 59% | 98% | 83% | 95% | 94% |
| Pyogenic brain abscess | 43% | 99% | 60% | 97% | 96% |
| Brain toxoplasmosis | 67% | 100% | 100% | 99% | 99% |
| Encephalitis | 50% | 97% | 55% | 96% | 94% |
| Vasculitis | 33% | 100% | 100% | 100% | 99% |
| Infarction | 67% | 99% | 86% | 98% | 97% |
| Non-viable brain tissue | 38% | 99% | 75% | 96% | 96% |
| Normal brain | 100% | 97% | 55% | 100% | 97% |

As indicated, MRS exhibited high specificity, NPV, and accuracy, approaching or exceeding 90% for all entities. Notably, MRS demonstrated promising results for diagnosing primary brain tumors, with sensitivity, specificity, PPV, NPV, and accuracy recorded at 74%, 97%, 92%, 88%, and 89%, respectively. The technique excelled in differentiating tumor recurrence from postoperative changes or gliosis in previously treated brain tumors, achieving values of 92%, 100%, 100%, 99%, and 99%, respectively. Furthermore, MRS exhibited high accuracies of 98% and 97% for diagnosing brain metastases and CNS lymphoma, both critical for differentiation from primary brain tumors.

Acceptable results were also observed for diagnosing demyelinating diseases, brain toxoplasmosis, encephalitis, brain infarction, and even normal brain tissue. However, the sensitivity and PPV of MRS for diagnosing pyogenic brain abscesses, CNS vasculitis, and non-viable brain tissue, such as gliosis and hematomas, were found to be below 50%, indicating that some of these pathologies were misdiagnosed by MRS as other entities.

For pyogenic brain abscesses, four cases went undiagnosed by MRS; one was misidentified as a primary brain tumor, while another was classified as non-viable tissue (gliosis or hematoma) by MRS, and the patterns of the two remaining cases were non-specific. For CNS vasculitis, two cases of Behçet’s disease were not diagnosed by MRS; one was reported as encephalitis and the other deemed non-specific.

Additionally, two cases of intracranial hematoma were misdiagnosed as brain abscesses by MRS; one exhibited a normal MRS pattern, and the other showed a non-specific pattern. One case of gliosis was also misinterpreted as a primary brain tumor by MRS. Figure 1 illustrates MRS findings for each pathology. For instance, MRS has correctly diagnosed 74% of primary brain tumors while 16% of cases of histopathology proven primary brain tumors are misdiagnosed by MRS as non-specific, 4% as encephalitis, and 2% as CNS lymphoma, demyelinating disease, and normal findings. Also, for pathology proven brain metastasis, MRS has correctly diagnosed 50% of cases, while 25% of them are misdiagnosed by MRS as primary brain tumors and 25% of them show non-specific spectroscopic pattern. Summary of all the included brain disorders are presented in the figure.

**Figure 1.**
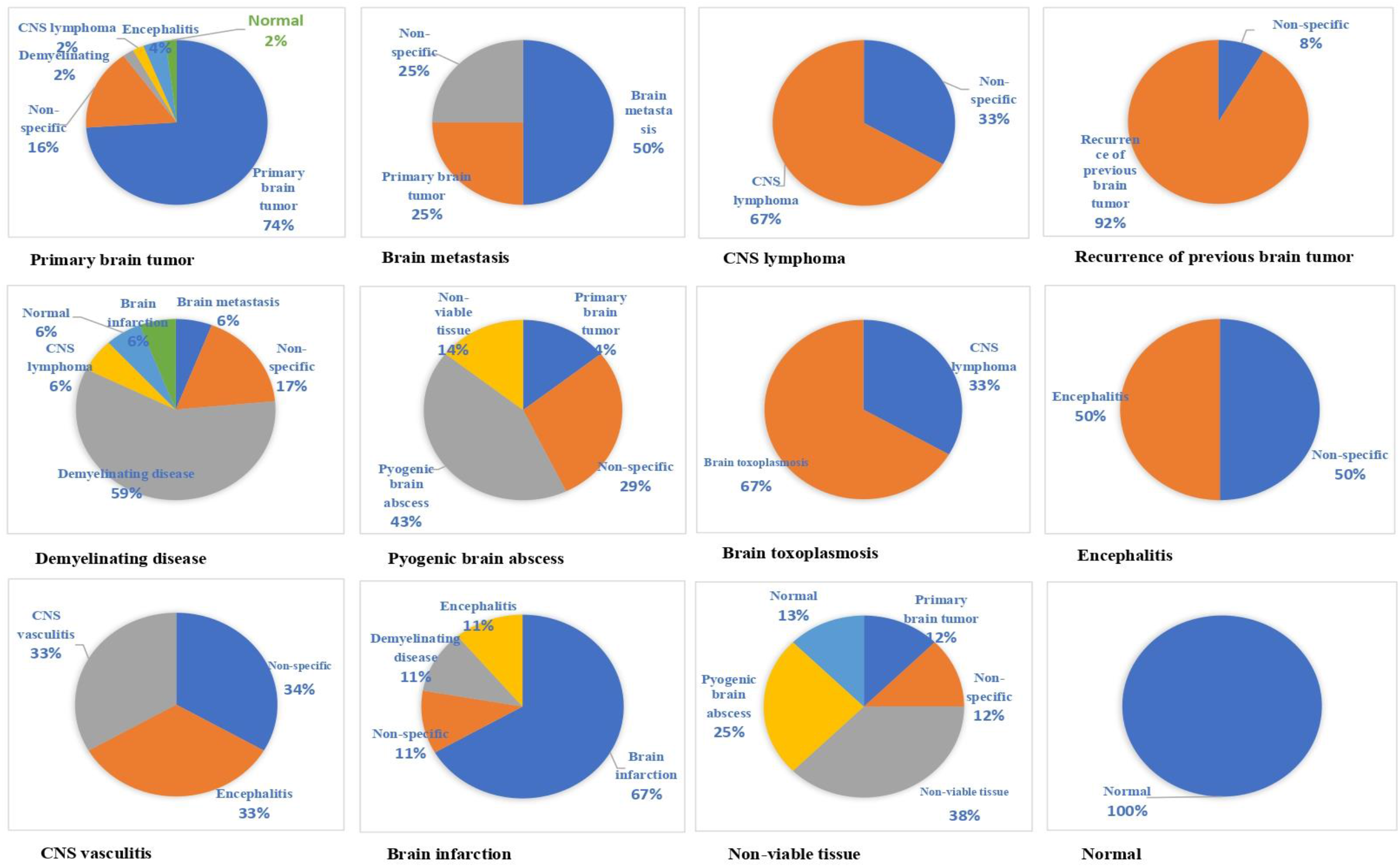
Summary of MRS final diagnosis for each pathology. For instance, MRS has correctly diagnosed 67% of CNS lymphoma while 33% of cases with further histopathology proven lymphoma show non-specific spectroscopic pattern not attributable to a specific disorder.

As noted earlier, 33 MRS studies did not yield a definitive diagnosis and provided multiple differentials based on observed patterns or stated that the patterns were non-specific without offering any differential diagnosis. Among these 33 studies, clinical data revealed that eight cases had a final clinical diagnosis of primary brain tumor, five were associated with encephalitis and inflammatory pathologies, three were attributed to demyelinating diseases, and the remaining cases corresponded to other pathologies (Figure 2).

**Figure 2.**
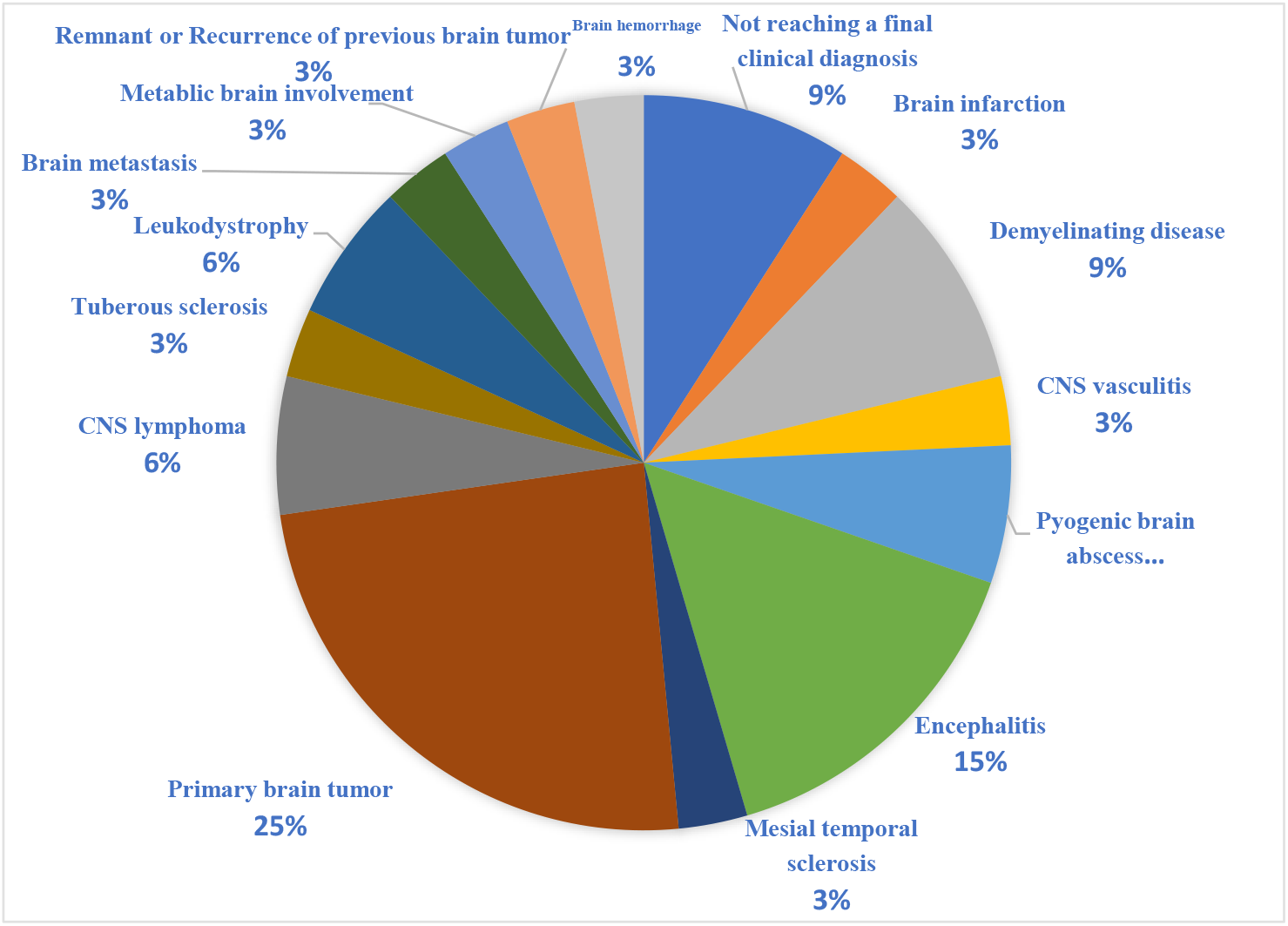
Summary of final clinical diagnosis of 33 cases with non-specific findings in MRS.

## DISCUSSION

This study analyzed 146 MRS investigations and found that MRS diagnoses were concordant with clinical diagnoses in 89 cases, resulting in a diagnostic accuracy of 61%. The analysis also demonstrated a specificity exceeding 90% across all included neurologic disorders, with overall diagnostic accuracy around or above 90%. The highest sensitivities were observed in the identification of normal brain tissue (100%) and the differentiation of tumor recurrence from post-treatment changes (92%). For neoplastic disorders, sensitivities were 74% for primary brain tumors, 67% for CNS lymphoma, and 50% for brain metastases. Non-neoplastic neurologic disorders showed sensitivities ranging from 33% to 67%, with the lowest scores associated with CNS vasculitis, non-viable brain tissue, and pyogenic brain abscesses. These findings support the reliable performance of MRS in diagnosing specific brain pathologies, primarily primary and secondary tumoral processes, while highlighting its limited ability to accurately diagnose some non-neoplastic conditions, consistent with previous literature (16–18).

While MRS demonstrated notable effectiveness—particularly in distinguishing tumoral processes such as primary brain tumors, metastases, CNS lymphoma, and tumor recurrence—it showed limited sensitivity for certain conditions, including pyogenic brain abscesses, hemorrhages, and CNS vasculitis (sensitivity <50%). Despite this, its specificity for these entities remained approximately 90%. These findings are consistent with existing literature, which documents characteristic spectroscopic patterns associated with various pathologies. Cases exhibiting these typical features are likely to be accurately classified with high specificity. However, the low sensitivity observed in some conditions suggests that atypical spectroscopic presentations—those deviating from classic patterns—may result in misdiagnosis. Additionally, many non-neoplastic neurological disorders can display a broad spectrum of spectroscopic patterns that differ from established profiles, potentially leading to incorrect diagnoses. Therefore, further studies are needed to explore and characterize these atypical spectroscopic patterns. Future research should aim to identify and understand such variations to improve diagnostic accuracy.

A review of the literature indicates that most prior studies have concentrated on MRS performance in diagnosing tumoral processes rather than other pathologies, with findings largely consistent with ours. For instance, a 2013 study compared the accuracy of MRS and conventional brain MRI in diagnosing various brain lesions. It reported an accuracy of 100% for high-grade gliomas, 66.7% for low-grade gliomas, and 40% for non-tumoral conditions such as benign lesions, demyelinating diseases, radiation necrosis, and infarcts. The overall accuracy was 83%, based on a small sample size of 23 patients (19). This study supports the notion that MRS reliably differentiates high-grade tumors and distinguishes between high-and low-grade gliomas, though its reliability for radiation necrosis and other non-tumoral pathologies was limited.

Few studies have included non-neoplastic neurological cases to assess their spectroscopic patterns. Therefore, this study is among the first to encompass a broad spectrum of such disorders. Additionally, the limited number of articles examining the spectroscopic features of these non-tumoral neurological conditions may explain the lower MRS scores observed for these pathologies, given the relative scarcity of research in this area

### Brain Tumoral Pathologies

MRS holds significant potential in the diagnostic evaluation of brain tumors. In MRS, both metastases and gliomas typically exhibit increased Cho and decreased NAA compared to normal white matter. Notably, lipids and macromolecules are more prominent in metastases than in glioblastomas. Spectroscopic analysis of peritumoral edema provides additional diagnostic clues, as gliomas often contain tumor cells within surrounding edema, resulting in elevated Cho/NAA and Cho/Creatine (Cr) ratios (7-9). An increased tumor-to-normal tissue ratio (Cho/NAA) in the peritumoral region correlates with tumor invasiveness and can help differentiate high-grade gliomas from metastases, which often show near-normal spectra in adjacent tissue (20).

Horska et al. reported an 84% accuracy in classifying 69 brain tumors using MRS. When combined with perfusion MRI, a sensitivity of 72.2% and specificity of 91.7% were achieved in differentiating tumors from non-neoplastic lesions (21). A large multicenter study demonstrated approximately 90% accuracy in distinguishing main adult tumor types, except for glioblastoma multiforme versus metastasis, where accuracy was 78%. For pediatric brain tumors, the accuracy reached 98%. Additional research showed that combining MRS with conventional MRI improved the prediction of tumor grade.

Our results align with existing literature, showing diagnostic accuracies of 89%, 98%, and 97% for primary brain tumors, metastases, and lymphomas, respectively, with specificities of 97%, 99%, and 98%. Sensitivities, however, were lower: 74%, 50%, and 67%, respectively. Notably, misdiagnosis of metastases as primary tumors contributed to the low sensitivity in detecting metastases (see Figure 1). These findings suggest that while MRS has considerable potential for differentiating brain tumor types, limitations remain. Importantly, MRS demonstrated high accuracy (92%) and specificity (100%) in distinguishing tumor recurrence from post-treatment changes, including radiation necrosis.

### Demyelinating Disease

The clinical utility of MRS in multiple sclerosis (MS) remains limited, despite insights into disease pathology and axonal damage assessment. Studies consistently report reduced NAA/Cr ratios in chronic MS plaques with occasionally elevated Cho/Cr ratios. Active inflammatory plaques often show elevated Cho/Cr and macromolecular signals—possibly reflecting myelin breakdown—alongside normal or decreased NAA/Cr ratios (4, 20, 22, 23).

MRS aids in differentiating tumefactive demyelinating lesions from neoplasms, though its diagnostic confidence remains debated. AlMadani et al. highlighted that, combined with clinical and conventional MRI data, MRS might serve as a non-invasive tool for delineating tumefactive lesions, but its standalone discriminative power is still uncertain (24).

In our study, MRS yielded a sensitivity of 59%, specificity of 98%, and overall accuracy of 94% for demyelinating plaques. The primary limitation was low sensitivity, mainly due to non-specific spectroscopic patterns in 17% of cases, often leading to misclassification as metastases, lymphoma, or infarct (Figure 1). These findings emphasize the need to recognize atypical MRS patterns associated with demyelination.

### CNS Infections

MRS can assist in confirming pyogenic abscesses and identifying causative organisms. Specific metabolite signals—succinate, acetate, alanine, leucine, isoleucine, and valine—are characteristic of pyogenic abscesses. MRS further differentiates tuberculomas with caseation from other lesions; tuberculous abscesses typically show lipid and lactate signals without cytosolic amino acids. Therefore, MRS plays a valuable role in diagnosing and managing focal brain infections (20, 25, 26)

However, a recent 2024 study demonstrated that MRS had limited accuracy in differentiating pyogenic abscesses from necrotic tumors, with sensitivity, specificity, and accuracy of 75%, 95.31%, and 38%, respectively (27). In our investigation, sensitivities were 43% for pyogenic abscesses, 67% for toxoplasmosis, and 50% for encephalitis; specificities, PPVs, NPVs, and overall accuracy were high (>90%). As shown in Figure 1, non-specific MRS patterns and atypical presentations contributed to diagnostic challenges, particularly in infections like toxoplasmosis, where differentiation from lymphoma is critical.

### Brain Infarction

While MRS is limited in acute stroke management due to logistical constraints, it offers promise beyond the hyperacute phase. It can assess neuronal damage and metabolic alterations, such as decreasing NAA, which correlates linearly with time post-ischemia (28, 29). In our study, MRS achieved sensitivity, specificity, and accuracy of 67%, 99%, and 97%, respectively, in diagnosing infarction. However, routine clinical use remains limited, and further validation is needed.

### CNS Vasculitis

CNS vasculitis exhibited low sensitivity in our analysis. Literature on MRS in vasculitis is sparse; one study noted differences in amino acid levels between primary CNS vasculitis and glioblastoma but lacked detailed statistical analysis (30). Further research is necessary to elucidate MRS’s diagnostic utility in vasculitis.

As mentioned, this study is the first to evaluate a broad spectrum of neurological pathologies with comprehensive statistical metrics. Future research should involve larger cohorts of patients with confirmed diagnoses who undergo MRS, enabling detailed characterization of spectroscopic patterns, including atypical features. Such efforts could refine diagnostic templates, improve accuracy, and enhance clinical decision-making. For instance, a sufficient number of confirmed CNS vasculitis cases, particularly those with the lowest sensitivity in our study, should be collected and subjected to retrospective MRS analysis to evaluate their spectroscopic features. Although we included a broad spectrum of disorders, the number of some cases was inadequate to reliably interpret their spectroscopic patterns.

Limitations of this study include the absence of histopathological confirmation for some cases, reliance on clinical diagnoses, and potential retrospective biases, as MRI and MRS data were accessed from picture archiving and communication system (PACS) with radiologists aware of MRI findings. Blinded prospective studies are warranted. Additionally, standardization of acquisition and analysis protocols— particularly voxel placement—is essential to improve reproducibility and diagnostic precision (31).

## CONCLUSION

In conclusion, while MRS demonstrates promising capabilities in diagnosing neurological conditions, its limitations in sensitivity for certain pathologies highlight the need for ongoing refinement. Future research should focus on enhancing MRS diagnostic accuracy, particularly for conditions with high misdiagnosis rates. This may involve developing standardized protocols, increasing training for interpreting MRS findings, and integrating advanced machine learning algorithms. Ultimately, MRS should be viewed as a complementary tool within a multimodal diagnostic framework that includes clinical evaluation and other imaging techniques.

## Data Availability

All data produced in the present study are available upon reasonable request to the authors

## Notes

### Competing Interest Statement

The authors have declared no competing interest.

### Funding Statement

This study did not receive any funding

### Author Declarations

Ethics committee/IRB of Shiraz University of Medical Sciences gave ethical approval for this work with the approval ID of IR.SUMS.MED.REC.1403.205.

## REFERENCES

1. Snyder J, Noujaim D, Mikkelsen T. Magnetic resonance spectroscopy. InHandbook of Neuro-Oncology Neuroimaging 2022 Jan 1 (pp. 385–394). Academic Press.

2. Zhu H, Barker PB. MR spectroscopy and spectroscopic imaging of the brain. Magnetic resonance neuroimaging: methods and protocols. 2011:203–26.

3. Buonocore MH, Maddock RJ. Magnetic resonance spectroscopy of the brain: a review of physical principles and technical methods. Reviews in the Neurosciences. 2015 Dec 1;26(6):609–32.

4. Sajja BR, Wolinsky JS, Narayana PA. Proton magnetic resonance spectroscopy in multiple sclerosis. Neuroimaging Clinics of North America. 2009 Feb 1;19(1):45–58.

5. Soares DP, Law M. Magnetic resonance spectroscopy of the brain: review of metabolites and clinical applications. Clinical radiology. 2009 Jan 1;64(1):12–21.

6. Weinberg BD, Kuruva M, Shim H, Mullins ME. Clinical applications of magnetic resonance spectroscopy in brain tumors: from diagnosis to treatment. Radiologic Clinics. 2021 May 1;59(3):349–62.

7. Pope WB. Brain metastases: neuroimaging. Handbook of clinical neurology. 2018 Jan 1;149:89–112.

8. Durmo F, Rydelius A, Baena SC, Askaner K, Lätt J, Bengzon J, Englund E, Chenevert TL, Björkman-Burtscher IM, Sundgren PC. Multivoxel 1H-MR spectroscopy biometrics for preoprerative differentiation between brain tumors. Tomography. 2018 Dec;4(4):172–81.

9. Chiang IC, Kuo YT, Lu CY, Yeung KW, Lin WC, Sheu FO, Liu GC. Distinction between high-grade gliomas and solitary metastases using peritumoral 3-T magnetic resonance spectroscopy, diffusion, and perfusion imagings. Neuroradiology. 2004 Aug;46:619–27.

10. Ikeguchi R, Shimizu Y, Abe K, Shimizu S, Maruyama T, Nitta M, Abe K, Kawamata T, Kitagawa K. Proton magnetic resonance spectroscopy differentiates tumefactive demyelinating lesions from gliomas. Multiple sclerosis and related disorders. 2018 Nov 1;26:77–84.

11. Saindane AM, Cha S, Law M, Xue X, Knopp EA, Zagzag D. Proton MR spectroscopy of tumefactive demyelinating lesions. American journal of neuroradiology. 2002 Sep 1;23(8):1378–86.

12. Kim SH, Chang KH, Song IC, Han MH, Kim HC, Kang HS, Han MC. Brain abscess and brain tumor: discrimination with in vivo H-1 MR spectroscopy. Radiology. 1997 Jul;204(1):239–45.

13. Pal D, Bhattacharyya A, Husain M, Prasad KN, Pandey CM, Gupta RK. In vivo proton MR spectroscopy evaluation of pyogenic brain abscesses: a report of 194 cases. American journal of neuroradiology. 2010 Feb 1;31(2):360–6.

14. Wilson M, Andronesi O, Barker PB, Bartha R, Bizzi A, Bolan PJ, Brindle KM, Choi IY, Cudalbu C, Dydak U, Emir UE. Methodological consensus on clinical proton MRS of the brain.

15. Zhu H, Barker PB. MR spectroscopy and spectroscopic imaging of the brain. Magnetic resonance neuroimaging: methods and protocols. 2010 Dec 15:203–26.

16. Möller-Hartmann W, Herminghaus S, Krings T, Marquardt G, Lanfermann H, Pilatus U, Zanella F. Clinical application of proton magnetic resonance spectroscopy in the diagnosis of intracranial mass lesions. Neuroradiology. 2002 May;44(5):371–81.

17. Alam MS, Sajjad Z, Hafeez S, Akhter W. Magnetic resonance spectroscopy in focal brain lesions. Journal of the Pakistan Medical Association. 2011;61(6):540.

18. Hollingworth W, Medina LS, Lenkinski RE, Shibata DK, Bernal B, Zurakowski D, Comstock B, Jarvik JG. A systematic literature review of magnetic resonance spectroscopy for the characterization of brain tumors. American journal of neuroradiology. 2006 Aug 1;27(7):1404–11.

19. Rao PJ, Jyoti R, Mews PJ, Desmond P, Khurana VG. Preoperative magnetic resonance spectroscopy improves diagnostic accuracy in a series of neurosurgical dilemmas. British journal of neurosurgery. 2013 Oct 1;27(5):646–53.

20. Öz G, Alger JR, Barker PB, Bartha R, Bizzi A, Boesch C, Bolan PJ, Brindle KM, Cudalbu C, Dinçer A, Dydak U. Clinical proton MR spectroscopy in central nervous system disorders. Radiology. 2014 Mar;270(3):658–79.

21. Horská A, Barker PB. Imaging of brain tumors: MR spectroscopy and metabolic imaging. Neuroimaging clinics of North America. 2010 Aug;20(3):293.

22. Richards TL. Proton MR spectroscopy in multiple sclerosis: value in establishing diagnosis, monitoring progression, and evaluating therapy. AJR. American journal of roentgenology. 1991 Nov;157(5):1073–8.

23. Davie CA, Barker GJ, Tofts PS, Hawkins CP, Brennan A, Miller DH, McDonald WI. Detection of myelin breakdown products by proton magnetic resonance spectroscopy. The Lancet. 1993 Mar 6;341(8845):630–1.

24. AlMadani A, AlShaffi SI, Abidzada A, Inshasi JS. Tumefactive demyelinating lesions in multiple sclerosis: role of magnetic resonance spectroscopy (MRS). ARCHIVOS DE MEDICINA. 2021;12(10):393.

25. Gupta RK. Magnetic resonance spectroscopy in intracranial infection. Clinical MR neuroimaging. New York: Cambridge University Press. 2010:426–51.

26. Chang KH, Song IC, Kim SH, Han MH, Kim HD, Seong SO, Jung HW, Han MC. In vivo single-voxel proton MR spectroscopy in intracranial cystic masses. American journal of neuroradiology. 1998 Mar 1;19(3):401–5.

27. Ashraf H, Ahsan K, Noor S, Kamran A, Ibrahim A, Arif A. Diagnostic Accuracy of Magnetic Resonance Spectroscopy and Diffusion-Weighted Mri in Differentiating Between Pyogenic Brain Abscesses and Necrotic Brain Tumors. Pakistan Armed Forces Medical Journal. 2024 Oct 31;74(5):1341.

28. van der Toorn A, Verheul HB, van der Sprenkel JW, Tulleken CA, Nicolay K. Changes in metabolites and tissue water status after focal ischemia in cat brain assessed with localized proton MR spectroscopy. Magnetic resonance in medicine. 1994 Dec;32(6):685–91.

29. Sager TN, Laursen H, Hansen AJ. Changes in N-acetyl-aspartate content during focal and global brain ischemia of the rat. Journal of Cerebral Blood Flow & Metabolism. 1995 Jul;15(4):639–46.

30. Kawazoe Y, Ohba S, Murayama K, Nakae S, Nishiyama Y, Abe M, Hasegawa M, Hirose Y. Contrast-enhanced magnetic resonance imaging, perfusion magnetic resonance imaging, and 1H-magnetic resonance spectroscopy distinguish primary central nervous system vasculitis from glioblastoma. World Neurosurgery. 2022 Feb 1;158:e820–8.

31. Ricci PE, Pitt A, Keller PJ, Coons SW, Heiserman JE. Effect of voxel position on single-voxel MR spectroscopy findings. American journal of neuroradiology. 2000 Feb 1;21(2):367–74.

